# Predicting IVF live birth probabilities using machine learning, center-specific models: validation results and potential benefits over national registry-based models

**DOI:** 10.1101/2024.06.20.24308970

**Authors:** Elizabeth T. Nguyen, Matthew G. Retzloff, L. April Gago, John E. Nichols, John F. Payne, Barry A. Ripps, Michael Opsahl, Jeremy Groll, Ronald Beesley, Gregory Neal, Jaye Adams, Lorie Nowak, Trevor Swanson, Xiaocong Chen, Mylene W. M. Yao

**Affiliations:** R&D Department, Univfy, Los Altos, CA, US; Fertility Center of San Antonio, San Antonio, TX, US; Gago Center for Fertility, Brighton, MI, US; Piedmont Reproductive Endocrinology Group, Greenville, SC, US; NewLIFE Fertility, Pensacola, FL, US; Poma Fertility, Kirkland, WA, US; SpringCreek Fertility, Dayton, OH, US

**Keywords:** live birth probability, IVF live birth prediction, artificial intelligence, machine learning, SART, fertility prognosis

## Abstract

Ongoing improvement of pretreatment live birth prognostication for in vitro fertilization (IVF) is critical for informing fertility patients’ treatment decisions, advocating for IVF coverage and supporting value-based IVF care. The US national registry Society for Assisted Reproductive Technology (SART) IVF live birth prediction (LBP) model (SART model) has been widely adopted for its prognostic support without external validation or utilization studies. We conducted a retrospective model validation study to compare the IVF LBP performance of machine learning, center-specific (MLCS) models versus the SART model in 6 unrelated US fertility centers using their respective center-specific test sets comprising an aggregate of 4,635 patients’ first-IVF cycle data. Compared to the SART model, MLCS2 showed higher median Precision Recall AUC at 0.75 (IQR 0.73, 0.77) vs. 0.69 (IQR 0.68, 0.71), p<0.05 and higher median F1 Score across LBP thresholds. Further, MLCS1 showed no evidence of data drift when validated using out-of-time test data from a later period. Reclassification analysis showed that MLCS2 models assigned more appropriate and higher IVF LBPs compared to the SART model, which underestimated patient prognoses (continuous net reclassification index: 18.3%, p<0.0001). Overall, MLCS2 and SART models assigned 30% of patients to differential prognostic groups, with MLCS2 assigning 26% of patients to a higher LBP category compared to the SART model. Importantly, MLCS2 models identified 11% of patients to have LBP ≥ 75%, whereas the SART model detected none. This group had a live birth rate of 81%. We recommend testing a larger sample of fertility centers to further evaluate MLCS model benefits and limitations.

## Introduction

Infertility, declared a medical disease and global health issue by the World Health Organization (WHO), is estimated to affect 107M-172M women or couples globally and over 10M in the US (1–14). Despite the proven safety and efficacy of assisted reproductive technology (ART), patients’ access to and utilization of ART continue to be met with barriers. Of the couples who could benefit from ART to have a family, only 5% was estimated to use it (4). Further, an estimated 10-50% of patients discontinued fertility care before achieving a live birth (15, *personal communications*).

Gaps at different levels of fertility care access included cost of treatment, stigma, emotional stress and uncertain probability of success (5–7, 14–15). (As in vitro fertilization (IVF) is the most common type of ART, the rest of the article will refer to IVF instead of ART.)

Our research has focused on improving IVF access and utilization for patients who have proactively and voluntarily sought fertility care at IVF centers. The potential contribution of artificial intelligence (AI)/machine learning (ML) to improve IVF pretreatment counseling warrants investigation on a broader scale, based on a reported 2-3 fold increase in IVF utilization among new patients across seven independent fertility centers in US and Canada (16). Providers have an important responsibility to offer validated, personalized and relevant prognostic counseling to educate patients about the potential benefits and limitations of IVF and to consider a course of IVF treatments to maximize the probability of having a baby (7, 17, 18). In addition, the cost of having an IVF baby is elusive for an individual patient because it depends heavily on the women’s or couple’s IVF live birth prognosis. IVF cost-to-live birth transparency is urgently needed for both patients and providers to support a very personal decision, whether for a medical or social/family need.

We previously reported the development and clinical usage of machine learning, center-specific (MLCS) IVF prognostic models to support provider-patient counseling across geographies (e.g. US, Canada, UK and EU) which varied in IVF payers including patients themselves, employers, health insurance plans and government (7, 16, 19–24). Geographically distributed, local delivery of fertility care helps to remove barriers in patient-centric care. However, patients’ clinical characteristics varied significantly across fertility centers and those inter-center variations were associated with differential IVF live birth outcomes (25). Therefore, patient-centric care includes providing patients with prognostic counseling that is relevant and reflects clinical characteristics of the local patient population.

Nonetheless, there is a widely held perception among providers that the national registry-based, center-agnostic IVF pretreatment prognostics model developed by the US Society for Assisted Reproductive Technology (SART model) “suffices” based on its usage of a large national dataset from 121,561 IVF cycles started in a two-year period (2014–2015) and its free online access, even though model validation using an external test set and clinical utilization studies have not been reported (26, 27). Further, recommendations were made to fertility centers in countries outside of US and UK to adapt local prognostic models by recalibrating the US SART and UK Human Fertilisation and Embryology Authority (HFEA) data-based models using local data, but Cai et al. showed that de novo MLCS model gave much improved model validation metrics (26, 28–30).

Validation of MLCS IVF pretreatment models using separate test sets have been reported and the design, validation and clinical usage considerations for IVF pretreatment prognostic model has been reviewed in-depth(7, 19, 20, 24, 28, 31, 32).

The above body of work led providers to ask 1) whether and how MLCS and SART model predictions differ for patients seeking fertility care in the US today, and 2) whether MLCS models are applicable to small-to-midsize US fertility centers. Indeed, a head-to-head comparison between the MLCS and SART pretreatment models for US centers has not been reported. Providers also want to know if “…the [MLCS] model has been validated for patients who received IVF after MLCS-based counseling?” Translated to ML and clinical research lingo, providers are asking “is there data drift causing previously validated models not to be applicable to patients doing IVF in a later time period?” Addressing these questions will help us to develop best practices for IVF live birth prognostic counseling, which is critical for advancing fertility care in the US and globally.

This retrospective cohort study aimed to compare the performance of the MLCS and SART pretreatment models for six unrelated small-to-midsize US fertility centers operating in 22 locations across 9 states in 4 US regions (West, Southeast, Southwest and Midwest) using their respective center-specific test sets comprising an aggregate of 4,635 patients’ first-IVF cycle data. This study’s primary outcome is a set of model performance metrics -- area-under-the-curve of the receiver operating characteristic curve (AUC-ROC), AUC improvement over age control, predictive power, precision, recall, F1 score, and precision-recall AUC -- used to evaluate MLCS and SART models (33). (Depending on the AMH availability of each cycle, the SART model with and without AMH as predictor would be used (26).) Live birth outcomes are primary outcomes for the prediction models being evaluated and are not primary outcomes of this study. We also addressed data drift, a scenario in which changes in patients, their characteristics, or treatment protocols cause a previously validated model not to be clinically applicable anymore (33, 34). To clarify, the AI techniques used and the problems addressed by this study are distinct from AI usage to improve embryo selection and its efficiency (35–37). Also, we did not use deep learning, foundational models or generative AI, so risks of “hallucination” do not apply (38–40).

## Results

Six centers participated in this study. Table 1 shows for each model validation, the MLCS model tested, time period of each data set, IVF volume range represented by the data set, data set usage (e.g. used for both training and testing or testing only), and the validation type (in-time or out-of-time).

**Table 1.** This table shows the time period, number of years and IVF volume represented by each data set matched against the MLCS model being tested. We also indicated whether (i) the dataset was used for both training and testing or testing only and (ii) the model validation was cross validation (CV) using in-time data or live model validation (LMV) using out-of-time data.

| Clinic | MLCS-based PreIVF model tested | Attributes of dataset used |  |  | Data use: both (train and test) or test only | Model Validation Type |  |
| --- | --- | --- | --- | --- | --- | --- | --- |
|  |  | Time period | Number of years | Number of IVF cycles (range) |  | CV or LMV | In-time or out-of-time |
| 916 | MLCS 1 | 2014-2016 | 3 | 501-1000 | both | CV | in-time |
|  | MLCS 2 | 2014-2020 | 7 | 1001-2000 | both | CV | in-time |
|  | MLCS 1 | 2017-2020 | 4 | 501-1000 | test only | LMV | out-of-time |
| 552 | MLCS 1 | 2014-2016 | 2.5 | 101-200 | both | CV | in-time |
|  | MLCS 2 | 2014-2020 | 7 | 301-500 | both | CV | in-time |
|  | MLCS 1 | 2016-2020 | 4.5 | 101-200 | test only | LMV | out-of-time |
| 635 | MLCS 1 | 2013-2016 | 4 | 501-1000 | both | CV | in-time |
|  | MLCS 2 | 2013-2020 | 8 | 1001-2000 | both | CV | in-time |
|  | MLCS 1 | 2017-2020 | 4 | 501-1000 | test only | LMV | out-of-time |
| 189 | MLCS 1 | 2014-2018 | 5 | 301-500 | both | CV | in-time |
|  | MLCS 2 | 2014-2020 | 7 | 501-1000 | both | CV | in-time |
|  | MLCS 1 | 2019-2020 | 2 | 201-300 | test only | LMV | out-of-time |
| 869 | MLCS 1 | 2014-2018 | 5 | 501-1000 | both | CV | in-time |
|  | MLCS 2 | 2016-2020 | 5 | 501-1000 | both | CV | in-time |
|  | MLCS 1 | 2019-2020 | 2 | 201-300 | test only | LMV | out-of-time |
| 395 | MLCS 1 | 2013-2018 | 6 | 501-1000 | both | CV | in-time |
|  | MLCS 2 | 2013-2021 | 9 | 1001-2000 | both | CV | in-time |
|  | MLCS 1 | 2019-2021 | 2 | 501-1000 | test only | LMV | out-of-time |

Abbreviations: MLCS1 = machine learning, center-specific model version 1; MLCS2 = machine learning, center-specific model version 2; CV = cross validation using in-time data; LMV = live model validation, a term we coined for external validation or the use of out-of-time data in model validation.

For each center, the initial, version 1 MLCS model (MLCS1) and the updated, version 2 MLCS model (MLCS2) cross validation results showed improved AUC and positive posterior log odds ratio compared to age (PLORA), indicating they were superior to their respective age control models. The median and interquartile range (IQR) of AUC, AUC improvement, and PLORA are summarized in Table 2. Next, we tested whether the AUC and PLORA of MLCS2 were improved over those of MLCS1. Across 6 centers, AUC was similar between MLCS1 & MLCS2, but PLORA of MLCS2 (23.9, IQR 10.2, 39.4) was improved over those of MLCS1 (7.2, IQR 3.6, 11.8), p<0.05. Therefore, the model update process (i.e. using a larger data set including more recent years of data) resulted in improved model performance (Table 2).

**Table 2.** This table shows the median and interquartile range (IQR) for cross validation and live model validation metrics -- AUC, AUC Improvement over Age-only model and PLORA -- for MLCS1 and MLCS2 models across 6 centers. See Methods for source of the age-only control model (61).

| Model | Validation | Model validation results: median and IQR |  |  |
| --- | --- | --- | --- | --- |
|  |  | AUC | AUC Improvement compared to age-only control | PLORA |
| MLCS 1 | CV, in-time | 0.66 (IQR = 0.61, 0.68) | 41.0% (IQR = 32.8%, 49.6%) | 7.2 (IQR = 3.6, 11.8) |
| MLCS 2 | CV, in-time | 0.67 (IQR = 0.66, 0.68) | 44.1% (IQR = 36.4%, 50.2%) | 23.9 (IQR = 10.2, 39.4) |
| MLCS 1 | LMV, out-of-time | 0.65 (IQR = 0.63, 0.66) | 7.6% (IQR = 5.2%, 8.7%) | 6.7 (IQR = 2.2, 12.0) |

Abbreviations: MLCS1 = machine learning, center-specific model version 1; MLCS2 = machine learning, center-specific model version 2; PLORA = posterior log odds ratio compared to age.

To test for the risk of data drift, we performed live model validation (LMV) testing on each center’s MLCS1 model using a center-specific, out-of-time data set. There was no detectable data drift based on observing comparable AUC and PLORA values between the LMV and cross validation (CV) results for MLCS1 model (Table 1).

MLCS2 and SART models were evaluated for each center using the modified, center-specific de novo model validation test sets, de novo model validation test sets 1 and 2 (DNMV1 and DNMV2), each comprising an aggregate of 4,645 and 4,421 unique patient-cycles across 6 centers. The overall rates of live birth labeling were 58.5% and 56.4% for DNMV1 and DNMV2, respectively.

Further, only ∼5% of patient-cycles did not have an AMH value and they were tested by the SART model formulae that did not require AMH predictor.

AUC and PLORA were not significantly different between the MLCS2 and SART models for either DNMV1 or DNMV2 (Table 2). Further evaluation was performed using the model metrics F1 Score (the harmonic mean of precision and recall) and Precision-Recall (PR) AUC, which are more sensitive than the AUC ROC in detecting improvements in predicting the positive class which is live birth prediction in the context of this study.

The median F1 Score was higher for MLCS2 compared to SART model across predicted live birth probability (LBP) thresholds sampled at ≥40%, ≥50%, ≥60%, ≥70%. For example, at the 50% LBP threshold, MLCS2 had a median F1 Score of 0.74 (IQR=0.72, 0.78) compared to 0.71 (IQR=0.68, 0.73) for SART using the DNMV1 test set. Similar findings were observed using the DNMV2 test set (Table 3).

**Table 3.** This table shows the median and interquartile range (IQR) for model cross validation metrics -- AUC ROC and PLORA -- measured by testing each center’s MLCS2 and SART models using each center’s de novo model validation test sets (DNMV1 and DNMV2) and using the SART 2020 age group-based live birth rate for each clinic as the age control “model”. *MLCS2 models showed significantly higher PR AUC score than SART, p<0.05.

| Test set for cross validation |  |  | Model validation results: median and IQR (in parenthesis) |  |  |  |
| --- | --- | --- | --- | --- | --- | --- |
| Test set | In-time or out-of-time data | Model | AUC ROC | PLORA | F1 score at 50% LBP threshold | PR AUC* |
| **DNMV1 test set (N=4,645)**<br>includes IVF cycles with clinical pregnancy outcomes | in-time | MLCS2 | 0.64<br>(0.62, 0.66) | 28.1<br>(15.2, 49.4) | 0.74<br>(0.72, 0.78) | 0.75<br>(0.73, 0.77) |
|  | out-of-time | SART | 0.65<br>(0.63, 0.66) | 22.5<br>(15.8, 46.5) | 0.71<br>(0.68, 0.73) | 0.69<br>(0.68, 0.71) |
| **DNMV2 test set (N=4,421)**<br>excludes IVF cycles with clinical pregnancy outcomes | in-time | MLCS2 | 0.64<br>(0.62, 0.66) | 23.5<br>(13.3, 33.9) | 0.74<br>(0.71, 0.75) | 0.73<br>(0.73, 0.75) |
|  | out-of-time | SART | 0.65<br>(0.62, 0.66) | 20.2<br>(14.9, 40.9) | 0.69<br>(0.67, 0.72) | 0.68<br>(0.66, 0.69) |

Abbreviations: AUC = area under the curve; ROC = receiver-operator curve analysis; PLORA = posterior log odds ratio compared to age control model; MLCS2 = machine learning, center-specific model version 2; DNMV = de novo model validation test set adapted to enable testing of MLCS2 and SART models; DNMV1 = the DNMV test set which included IVF cycles with clinical pregnancy outcomes; DNMV2 = the DNMV test set which excluded IVF cycles with clinical pregnancy outcomes; PR AUC = Precision-Recall AUC

PR AUC was also significantly higher for MLCS2 using DNMV1. The median PR AUC was 0.75 (IQR=0.73, 0.77) for MLCS2 and 0.69 (IQR=0.68, 0.71) for SART across the 6 centers, p<0.05 (Table 2). The findings were similar when tested using DNMV2 test set, p<0.05 (Table 3). At these six centers, assessing recall at the LBP threshold of ≥ 50%, MLCS2 models can identify ∼84% of patients who would go on to have IVF live births, while the SART model can only identify ∼75%. In other words, the use of SART model would have resulted in 9% of patients missing a good prognosis of ≥50% LBP. While overall precision was comparable between MLCS2 and SART, across all precision rates, MLCS2 models showed higher rates of recall and more cycles with higher IVF live birth probabilities across six centers (Figure 1A). The higher recall rates of MLCS2 are consistent with MLCS2 model generating an LBP distribution that is shifted to the right (i.e. having more patients with higher LBPs) compared to SART model (Figure 1B). Similar findings were observed for DNMV2.

**Figure 1.**
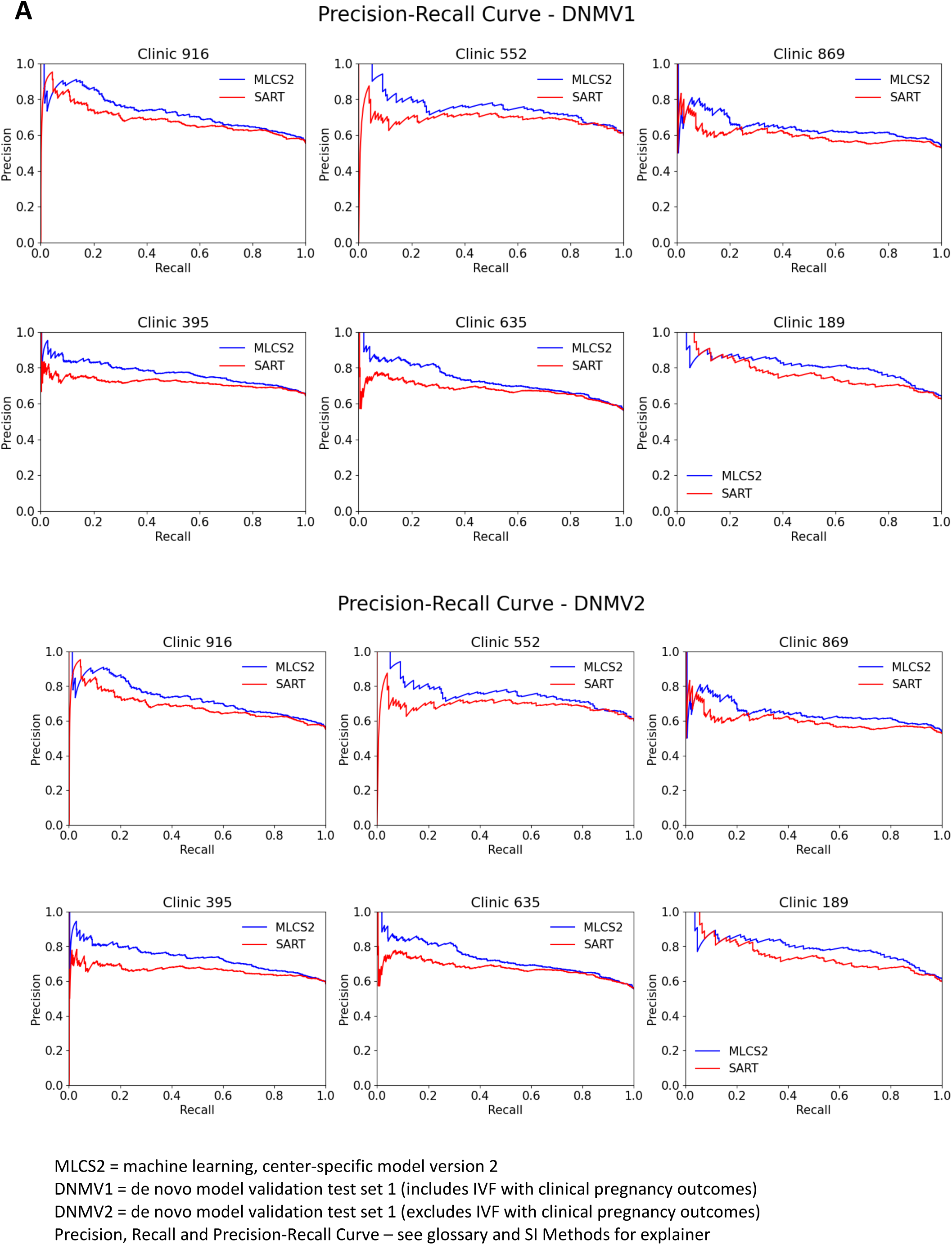
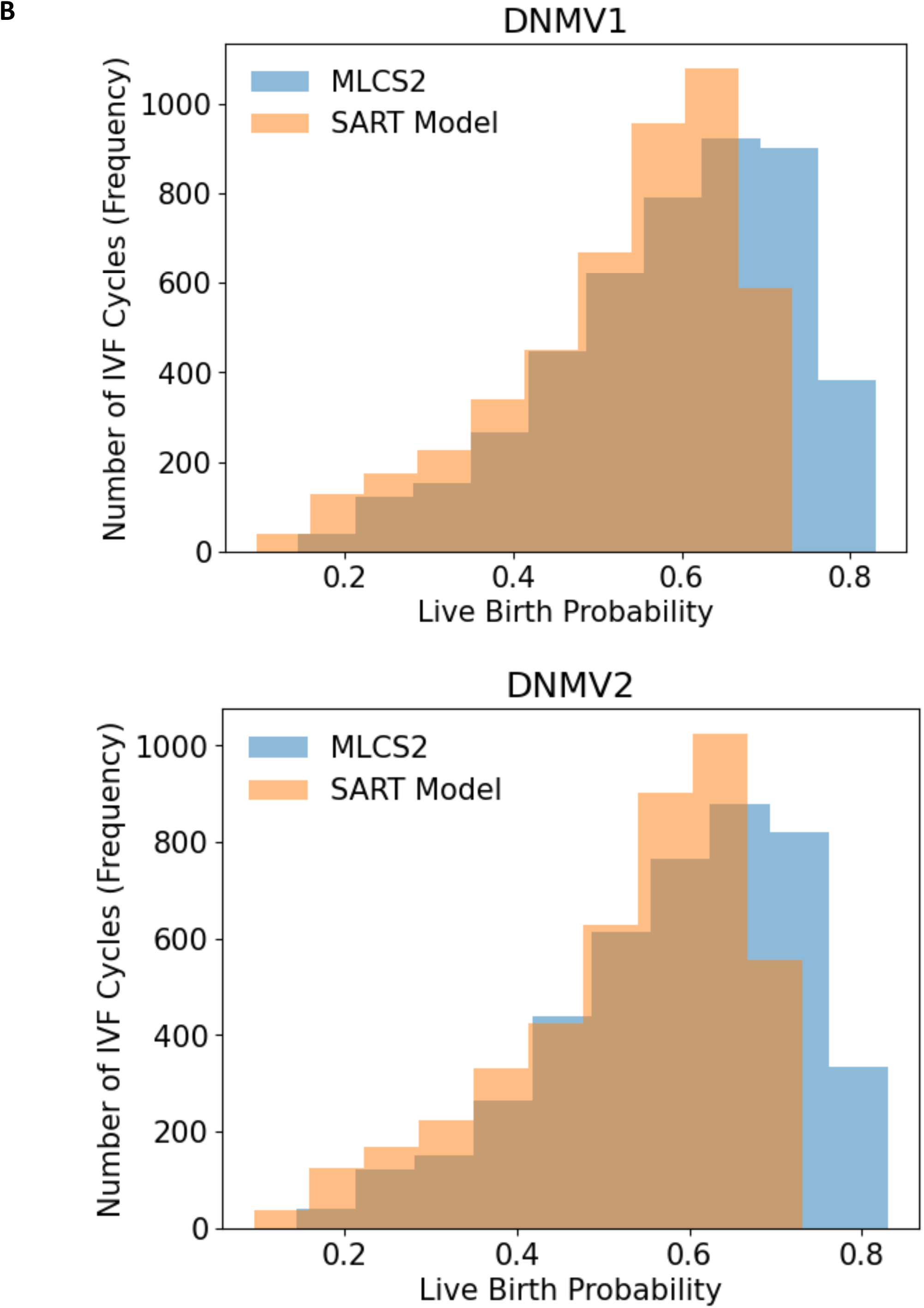
Comparison of MLCS2 and SART models using (A) Precision-Recall curves for each of the 6 clinics using each center’s de novo model validation test sets, DNMV1 and DNMV2; (B) frequency distributions of live birth probabilities using MLCS2 and the SART models for DNMV1 and DNMV2 test sets each comprising data from 6 centers’ test sets in aggregate.

Although the PR AUC, F1 Score and recall rates sufficed to show that the MLCS2 models provide more appropriate LBPs compared to the SART model, those model metrics were not intuitive for clinicians and patients. To facilitate communications with clinicians, we created a 4x4 reclassification table for the DNMV1 test set to show the concordance and discordance of LBPs made by MLCS2 and SART models in a practical, clinical context (see Methods and Table 4). (Note: Analyses related to model validation, LBPs and live birth rates were all necessarily performed at the group level, not at the individual patient level because it would not be possible or scientifically feasible to evaluate those measures on individual patients.)

**Table 4.** Reclassification table comparing IVF live birth prediction (LBP) models MLCS2 and SART across centers for (A) DNMV1 and (B) DNMV2. B. The same reclassification procedure (above) was applied to the DNMV2 test set.

|  |  | MLCS2 Model Prognostic Categories (LBP range) |  |  |  | Subtotal |
| --- | --- | --- | --- | --- | --- | --- |
|  |  | Low LBP<br>< 25% | Medium LBP<br>25% to 49.9% | High LBP<br>50% to 74.9% | Very High LBP<br>≥ 75% |  |
| SART<br>Model<br>Prognostic<br>Categories<br>(LBP<br>range) | Low LBP<br>< 25% | N = 79<br>LBR = 9% | N = 168<br>LBR = 33% | N = 2<br>LBR = 50% | N = 0 | N = 249<br>LBR = 26% |
|  | Medium LBP<br>25% to 49.9% | N = 12<br>LBR = 0% | N = 735<br>LBR = 38% | N=566<br>LBR = 61% | N = 7<br>LBR = 100% | N = 1320<br>LBR = 48% |
|  | High LBP<br>50% to 74.9% | N = 0 | N = 144<br>LBR = 37% | N=2445<br>LBR = 64% | N = 487<br>LBR = 80% | N = 3076<br>LBR = 66% |
|  | Very High LBP<br>≥ 75% | N = 0 | N = 0 | N = 0 | N = 0 | N = 0 |
|  | Subtotal | N = 91<br>LBR = 8% | N = 1047<br>LBR = 37% | N = 3013<br>LBR = 64% | N = 494<br>LBR = 81% | N = 4645<br>LBR = 59% |

**Table 4 (Cont'd.)** B. The same reclassification procedure (above) was applied to the DNMV2 test set.
|  |  | MLCS2 Model Prognostic Categories (LBP range) |  |  |  | Subtotal |
| --- | --- | --- | --- | --- | --- | --- |
|  |  | Low LBP<br>< 25% | Medium LBP<br>25% to 49.9% | High LBP<br>50% to 74.9% | Very High LBP<br>≥ 75% |  |
| SART<br>Model<br>Prognostic<br>Categories<br>(LBP<br>range) | Low LBP<br>< 25% | N = 78<br>LBR = 8% | N = 163<br>LBR = 31% | N = 1<br>LBR = 0% | N = 0 | N = 242<br>LBR = 24% |
|  | Medium LBP<br>25% to 49.9% | N = 12<br>LBR = 0% | N = 726<br>LBR = 37% | N=525<br>LBR = 58% | N = 7<br>LBR = 100% | N = 1270<br>LBR = 46% |
|  | High LBP<br>50% to 74.9% | N = 0 | N = 143<br>LBR = 36% | N=2336<br>LBR = 63% | N = 430<br>LBR = 78% | N = 2909<br>LBR = 64% |
|  | Very High LBP<br>≥ 75% | N = 0 | N = 0 | N = 0 | N = 0 | N = 0 |
|  | Subtotal | N = 90<br>LBR = 7% | N = 1032<br>LBR = 36% | N = 2862<br>LBR = 62% | N = 437<br>LBR = 78% | N = 4421<br>LBR = 56% |

Overall, 70% (3259 of 4645) of patients had concordant LBPs between SART and MLCS2 models (Table 4, green cells), while 30% (1386 of 4645) of the patients had discordant LBPs (Table 4, peach or blue cells) -- meaning, MLCS2 and SART models placed 1386 patients into different prognostic categories. Importantly, for the 30% of patients with discordant LBP prognostic categories, each group’s live birth rate aligned with MLCS2 model predictions whether MLCS2 assigned a higher or lower prognostic category than SART. Of the patients with discordant LBPs, 89% (1230 of 1386) were given higher LBPs by MLCS2 (Table 4, blue cells) and 11% (156 of 1386) were given lower LBPs by MLCS2 (Table 4, peach cells) compared to SART model.

Patient groups (in blue) that got moved up to a higher prognostic category by MLCS2 showed live birth rates that were in the range predicted by MLCS2 models. For example, of the patients that the SART model assigned to Low LBP (N=249), Medium LBP (N=1320) and High LBP (N=3076), 67%, 43% and 16%, respectively, were upgraded to a higher prognostic group by MLCS2 models. Further, these “upgraded” patient groups had live birth rates (LBR) that matched with the expected LBP range: patients upgraded from Low LBP to Medium LBP had 33% LBR; patients upgraded from Medium LBP to High LBP had 61% LBR; and patients upgraded from High LBP to Very High LBP had LBR 80%. (Table 4.)

Importantly, no patients received LBP ≥ 75% from the SART model whereas 11% of patients received LBP ≥ 75% from the MLCS2 models and the live birth rate for patients in this Very High LBP prognostic group was 81%. In summary, MLCS2 assigned 26% (1230 of 4645) of patients to a higher LBP category compared to the SART model. SART-LBPs underestimated live birth rates for each of those discordant groups across the spectrum of prognostic categories.

The converse was also observed: 12 of 1320 (1%) patients assigned to Medium LBP and 144 of 3076 (5%) patients assigned to High LBP by SART were downgraded to Low LBP and Medium LBP categories by MLCS2 models, respectively, yielding 0% LBR in the Low LBP group and 37% LBR in the Medium LBP group. Therefore, across all discordant groups, MLCS2 gave more appropriate LBPs compared to the SART model based on alignment of each group’s LBR to the expected LBP range for the MLCS2-associated prognostic category. Similar results were obtained from the DNMV2 test set.

Using continuous net reclassification index (NRI), compared with unsuccessful patients, patients with a live birth outcome were 18.3% (95% CI 13.3%, 23.2%) more likely to be given a higher LBP with MLCS2 compared to SART when tested using the DNMV1 (p<0.001). Similar findings were obtained using the DNMV2. Although, the differential prognoses affecting 30% of patients were not a measure of the models, they help to contextualize the improved model metrics in PR AUC, F1 Score and Recall observed for the MLCS2 over the SART models.

## Discussion

This study compared individual MLCS models and the SART model for pretreatment IVF live birth prognostics for six unrelated, geographically distributed US fertility centers that reported to the SART registry. The retrospective study design was appropriate because the prognostic models were previously trained, tested, and already in clinical usage, and evaluation of the models’ technical performance were not biased by the retrospective design. MLCS model validation was performed prior to clinical usage and the deployed MLCS models were tested using an out-of-time test set, coined live model validation, LMV. In addition to validating the models, those results also indicated that there was no detectable data drift.

We took the pragmatic realist approach to address “how do the MLCS and SART model predictions differ for the patients seeking care today?” The MLCS2 models performed better than the SART model in predicting the positive class (i.e. live birth prediction), as indicated by the PR AUC, F1 score and Recall (33). Further, using the 4 x 4 reclassification table to define concordance and discordance between MLCS2 and SART model LBPs provided clinically intuitive interpretation of the results. Overall, 30% of all patients in the DMNV1 test set showed discordance between MLCS2 and SART model LBPs, and of those 89% of patients were placed in lower prognostic groups by SART whereas 11% of patients were placed in higher prognostic groups by SART. All discordant groups received more appropriate LBPs from MLCS2 models than from the SART model, whether the discordance stemmed from under- or overestimation of LBPs by the SART model. Further, the under- and overestimation by SART affected patients with LBPs across the spectrum from Low LBP to Very High LBP were affected.

Why are these results important? First, patients and providers should be aware of the best available source of IVF live birth prognosis to inform patients’ decision-making. No additional justification should be needed. Patients who have very poor prognosis deserve to know their specific treatment limitations to inform funding IVF and subjecting themselves to physical intervention, whereas patients with good or excellent prognosis should not be discouraged by an underestimation of prognoses, which may deter or delay IVF, potentially resulting in not having a family. Further, in countries that allow the use of donor egg IVF, patients with poor prognosis may choose to use donor eggs for a higher probability of success.

The improved F1 Score and PR AUC model metrics were reflected by a high percentage of patients placed into groups with more appropriate and higher LBPs given by MLCS2 and underestimation of LBPs by SART. These findings can directly translate to support IVF pricing to the lower cost per IVF baby, as explained further in Yao et al. (24). This type of IVF pricing is already offered to self-pay patients or consumers, but it can be expanded and adapted for enterprise payers such as health plans and employers. For context, the cost per IVF baby is a significant barrier to IVF access, and in most US states, there is no mandated IVF coverage by health insurance plans (41, 42). Many fertility centers offer shared risk or refund programs to self-pay patients, with the goal of increasing the feasibility of doing a course of several IVF treatments. However, if the actuarial-like models backing these shared risk programs have suboptimal F1 Score, PR AUC and Recall, more patients may be deemed ineligible to enter the program. Therefore, achieving high precision and recall would alleviate the financial barrier while maximizing live birth outcomes for many patients (24).

The MLCS modeling framework enables providers to offer true value-based IVF care at scale, through transparent IVF pricing and live birth outcomes specific to each fertility center.

Prediction models that drive value-based IVF care can also support provider-patient counseling, an important part of patient-centric care. Most importantly, transparency can be achieved by using the same IVF live birth prediction model to support both value-based care and provider-patient counseling. The SART model, in the format of a free online calculator, presumably serves as an educational tool that encourages patients to seek care. However, at the point when patients have completed their diagnostic workup and are being counseled by providers about the benefits and limitations of IVF, patients are interested to know their IVF live birth probabilities at that particular center.

The MLCS design aimed to strengthen the patient-provider relationship by prioritizing patients’ top concern: “Is this IVF success prediction based on your center’s own data?” Whereas the MLCS live birth prediction directly responds to that question, the SART online calculator was not designed to address that concern. The SART online calculator’s disclaimer states, “The estimates are based on the data we have available and may not be representative of your specific experience…Please speak with your doctor about your specific treatment plan and potential for success.”

Like many innovative products used in healthcare, the MLCS-based counseling report has been developed and is sold by Univfy Inc., for-profit company. Currently, Univfy charges a fee to fertility centers for the center-specific implementation of the MLCS-based counseling model. Fertility centers using the MLCS-based report have independently chosen to fund the cost of these reports as a complimentary service to their patients. Therefore, patients are currently not bearing the cost of this technology and it is also equitably priced based on local IVF pricing and IVF volume. The authors of this study believe it is a public service to share this research study’s findings as we expect providers, researchers and patients would appreciate knowing about the capabilities of MLCS models. We leave it to stakeholders of the free market -- comprising patients, providers, private equity investors, biopharma, employers, health insurance plans and benefits companies -- to choose performance over freemium based on their patients’ and providers’ needs.

MLCS-based counseling reports may also improve patient-centric care by streamlining provider-patient flow and clinical workflow. In the context of clinical workflow, MLCS models can support diverse healthcare providers -- such as advanced practice providers (APPs), nurse practitioners and general obstetrician-gynecologists -- in performing patient counseling, further improving scalability and accessibility of IVF treatments (43, 44). MLCS models and their associated counseling reports have also been developed to support other ART clinical counseling scenarios -- e.g. after one or more failed IVF cycles, prior to egg freezing, whether to use their own eggs or eggs from a donor -- which included addressing patients with a poor prognosis and delivering prognosis with compassion (7, 19, 21–24, 45).

Having discussed the application and scope of real-world benefits of MLCS models -- including patient-centric care and lowering the cost per IVF baby -- we now turn to the technical aspects affecting the differential performance of MLCS and SART models. Considering the SART model used 121,561 IVF cycles whereas the MLCS2 models used a median dataset size of 1163 IVF cycles (IQR 658-1662 IVF cycles) to achieve comparable ROC AUC and improved PR AUC and F1 score, the MLCS approach is 200x more data efficient (26). Here, we hypothesized several factors to be driving the improved metrics: 1) The greater number of consecutive years covered by the MLCS data sets allowed for more freeze-all cycles to generate outcomes that reflect more realistic and higher live birth probabilities. 2) The lower-than-expected SART model ROC AUCs for the DNMV datasets may have resulted from the under 40 age limit of this study, whereas the original report of SART model training included age up to 50 years of age (24, 33). Inclusion of older patients can artificially increase the ROC AUC and that issue is discussed in-depth in a separate review (33). 3) MLCS enables greater flexibility in the number and scope of clinical predictors that can be tested for use in the prediction model to capture greater inter-patient differences and dynamic range of model prediction (7, 24-25, SI Figure 1). 4) Protocolization, maintenance of data processing and modeling pipelines, quality assurance, expert human supervision and most importantly, close collaboration with providers and centers’ operational teams likely contributed to the quality and validation of MLCS models (7, 24). As the use of ML gains maturity in healthcare, the emphasis shifts to delivering highly scalable, secured pipelines for model pre-processing, model training and model deployment (46). The quality control and considerations used from design to deployment specific to the production of IVF pretreatment live birth prediction models for use at the point-of-care are reviewed by Yao et al, 2024 (24).

Having outlined the likely reasons for improvement, we recommend viewing the differences between the MLCS2 and SART models in total. Instead of dissecting older models and datasets, we recommend to focus collaborative research efforts to study model metrics on a wider range of fertility centers -- larger centers, academic centers, centers in IVF coverage-mandated states and publicly funded IVF -- to learn the potential benefits and limitations of the use of center agnostic and center-specific multicenter models in comparison with MLCS models, for the prediction of IVF live birth probability in different clinical settings.

More broadly, the highly scaled MLCS framework -- creating MLCS models for many centers with collaboration with providers and quality control for the modeling -- can be used to advance reproductive research. As local specificity is crucial in solving health inequities in fertility care and IVF, which have remained significant and largely unsolved (47, 48), the MLCS framework can be applied to dissect the contribution of social determinants of health to fertility care utilization, access to care and IVF live birth outcomes. In addition, the MLCS framework can be applied to support the evaluation of add-ons, which aimed to improve IVF live birth outcomes but may have inconclusive or controversial clinical results. The acceptance of add-on procedures and the evaluation of each type of add-on have been challenging due to a combination of factors including the heterogeneity of the studies and patient populations (49). Those challenges relate to evaluating IVF patients as a whole and not having locally validated prognostic groups. Ultimately, amassing insights from many MLCS models is expected to advance precision medicine in IVF with cost-efficiency and capabilities to evaluate new diagnostic and therapeutic interventions.

To summarize, we have established a globally applicable framework for MLCS modeling of IVF live birth prediction to inform locally relevant patient-provider counseling, clinical workflow and value-based IVF care to lower the cost per IVF baby. Collectively, improvements in these areas are expected to contribute towards making IVF treatment accessible to more patients with a resulting increase in the families built and number of singleton babies born. We believe multicenter collaboration and collaboration between public and private sectors will accelerate and scale research tackling crucial questions related to racial disparities in IVF, molecular mechanisms of clinical infertility and IVF success, and ways to expand access to IVF care. We hope this study will help to advance reproductive medicine beyond dichotomies of multicenter versus center-specific or ML versus non-ML prediction models. Ultimately, the multicenter scaling of a machine learning, localized approach is expected to maximize benefit to people wanting a family while addressing health inequities and patient needs, de-risking IVF costs and advancing precision medicine in reproductive health.

## Methods

### Research data sources, de-identified data sets and prior reporting of methods

De-identified IVF treatment clinical variables and outcomes data previously linked and processed as part of Univfy client services were anonymized and entered into Univfy research database as per research protocol. The original data sources included electronic medical record (EMR) and SART CORS, the US national registry database managed by SART (50). Univfy Inc. submitted research protocol to institutional review board (IRB) which designated the exempt status for our research protocol, which used anonymized data.

Briefly, definitions of IVF treatments, live birth and methods used for data collection, exclusion criteria, use of center-specific variables and MLCS model life cycle and evaluation steps including model training and testing, gradient boosted machines (GBM) on the Bernoulli distribution, and the use of ROC-AUC, AUC improvement over age control model (”AUC improvement”) and posterior log of odds ratio (PLORA) were substantially as previously reported and are summarized here for ease of reference (7, 19, 20, 24, 51, Figure 2). Advantages of machine learning in general and GBM specifically over conventional methods such as logistic regression were reviewed previously (7, 24, 51). The first page of a sample provider-patient IVF pretreatment counselling report based on MLCS, live birth probability prediction model, commercially known as the Univfy® PreIVF Report, is available at https://www.univfy.com/research (52). In the MLCS models and throughout this article, an IVF cycle is defined as gonadotropin ovarian stimulation cycle with the intention of retrieving oocytes for IVF or ICSI for embryo culture and blastocyst transfer with or without PGT-A or freeze-all.

**Figure 2.**
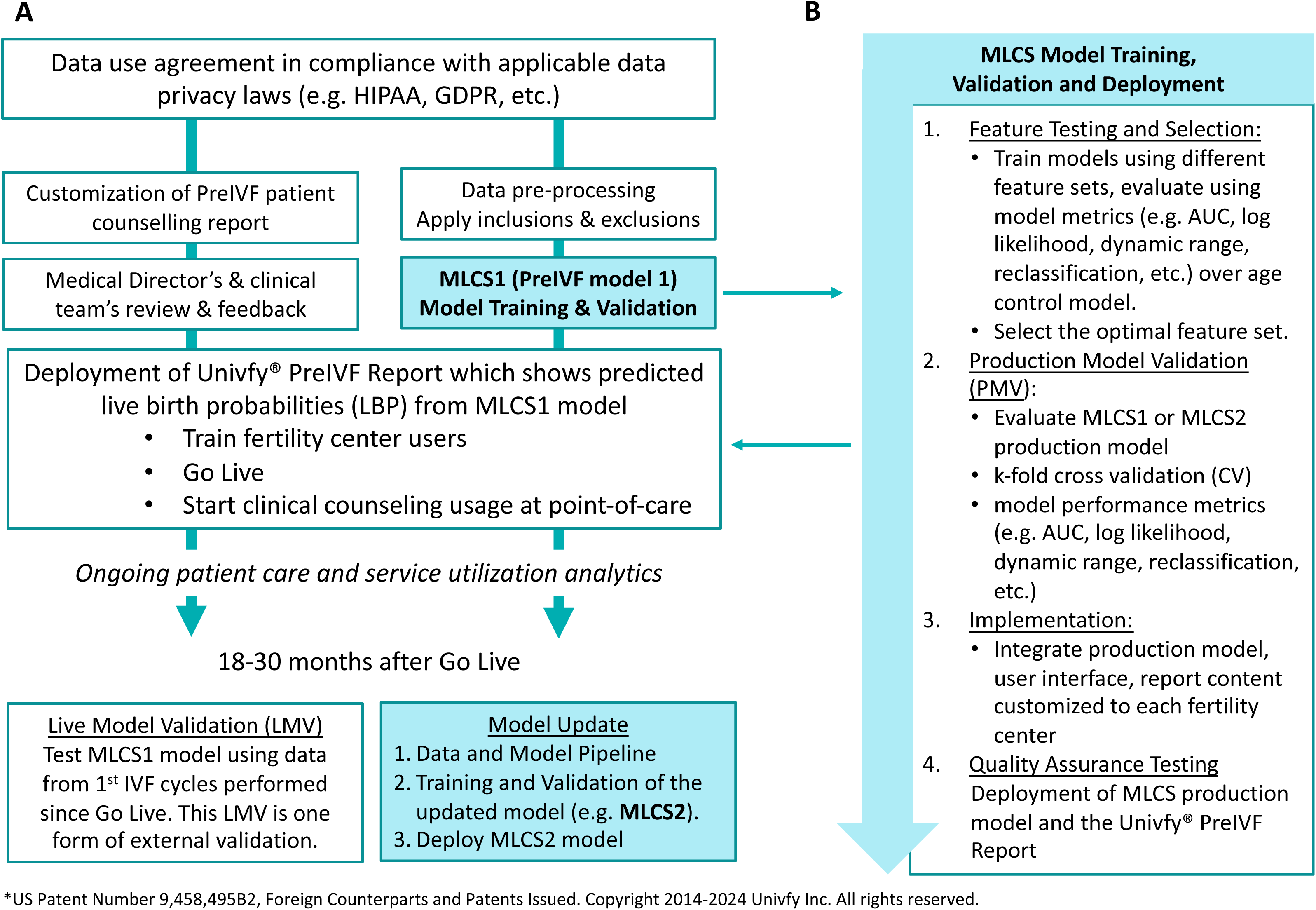
The development-to-deployment life cycle of the machine learning-based, center-specific (MLCS), prognostic model for use at point-of-care to support patient counseling*. (A) The MLCS-based, PreIVF model (MLCS model) product life cycle comprises the steps of data pre-processing, model training and validation, deployment and post-deployment validation (or live model validation). MLCS1, MLCS2, etc. indicates that each MLCS model will be replaced by an updated MLCS model trained and tested with a more recent data set which may also become cumulatively larger. (B) Model pipeline supports feature testing, model training, validation analysis, deployment to production and quality testing. “MLCS” is used generically to indicate the steps used for MLCS1, MLCS2 or any subsequent updates of MLCS model for a particular fertility center.

### Cross validation (CV) and model metrics

Our standard model evaluation procedure required k-fold cross validation on an in-time test set (the test and training data sources were contemporaneous) to compute the ROC-AUC, AUC improvement over age control model (“AUC improvement”) and the posterior log odds ratio compared to age control model (PLORA) as previously reported. In layman terms, PLORA describes “given a certain LBP prediction, how much more likely will the MLCS model be correct compared to age control?” PLORA, expressed in the log scale with log base e; non-statisticians may prefer to translate to linear scale (e^PLORA^) for more intuitive understanding (7, 19, 20, 24, 51, 53, Figure 2). CV of both MLCS1 and MLCS2 was reported using median and interquartile range (IQR) across 6 centers for ROC-AUC, ROC-AUC improvement and PLORA.

Briefly, a model with higher Precision (aka positive predictive value, PPV) is more likely to be correct when predicting a successful IVF live birth outcome; a model with higher Recall (aka sensitivity or true positive rate, TPR) would correctly identify a larger proportion of successful IVF live birth outcomes. F1 score measures the harmonic mean of Precision and Recall such that a high F1 score describes a model with both high Precision and high Recall at a particular live birth probability threshold (24, 33, 54). See *” Creating center-specific, de novo model validation test sets (DNMV1 and DNMV2) to enable statistical analyses of metrics of the MLCS2 and SART models”* below for detailed methods used to create and analyze test sets for model comparison.

### Data used for model training and testing, inclusion and exclusion criteria

Consecutive years of data within the 2013-2022 period were used for model training and testing performed for each center independently of the others. Each center’s Univfy report usage period started in 2016-2019 with data collection ending in 2020-2022 (Table 1). To assess the risk of data drift, we performed post-deployment, live model validation (LMV) per center, using an out-of-time test set from a time period following and exclusive from the MLCS1 training and test data (24, 34, 55, SI Figure 1). Using a larger, more recent, historical data set, each center’s first model (MLCS1) was replaced by an updated model (MLCS2) using the same MLCS model life cycle and evaluation (SI Figure 1). The MLCS2 models were in clinical use at the time of writing this article.

Inclusion criteria for model training and testing were: 1) IVF cycles started, 2) IVF cycles using/ intending to use the patient’s own eggs and uterus. Exclusion criteria were: FETs not linked to an original IVF ovarian stimulation cycle within that dataset; cancellations for reasons unrelated to IVF (e.g. covid, personal reasons, etc.); the use of donor egg, gestational carrier, embryo donation, egg freezing or fertility preservation for any reason; cycles without the female patient’s age or outcome; cycles with age 42 and up; freeze-all cycles that have not had any FETs and batched cycles; PGT-M usage. IVF cycles that resulted in cancellation of egg retrieval, no blastocyst or euploid blastocyst available for transfer or PGT-A usage were not excluded.

### IVF labeling criteria in the data used for MLCS training and testing

IVF cycles were labeled as having “no live birth outcome” if they resulted in cancellation at any point due to reasons related to the IVF cycle (e.g. poor ovarian response, thin endometrium, no viable oocytes, no fertilization, no blastocyst for transfer or no euploid blastocyst, etc.). In the context of MLCS models, an IVF cycle has achieved a live birth outcome if at least one live birth or clinical ongoing pregnancy were documented from one or more fresh and/or frozen embryo transfer(s) using the number of blastocyst(s) according to ASRM guidelines (56, 57).

To be clear, while MLCS and SART pretreatment models had live birth outcomes as primary outcomes for model prediction, live birth outcome itself is not the primary outcome in this study. Rather, this study’s primary outcome is to evaluate MLCS and SART models using model performance metrics. However, comparison of models’ metrics requires providing methods used to process data, train and test MLCS pretreatment models.

### Model predictors

The model predictors used by each center’s MLCS2 model and their relative importance vary across centers, despite drawing from a similar set of clinical variables (27-29, SI Figure 1). The SART pretreatment model predictors were either also used by MLCS2 models or were determined to have no non-redundant predictive impact based on other predictors used by the MLCS2 models. The MLCS2 models used additional predictors not used by the SART model even though they were recorded in SART-CORS (24, 26, SI Figure 1).

MLCS enables greater flexibility in the number and scope of clinical predictors that can be tested for use in the prediction model to capture greater inter-patient differences. Although the MLCS and SART model training sets were comparable in including female patient’s age, BMI, clinical diagnoses, and reproductive history, the MLCS models used one or more ovarian reserve tests (e.g. AMH, D3 FSH, or AFC) reflecting each center’s practice without being affected by inter-center laboratory differences irrelevant to each center. However, AMH value was available in ∼95% of cycles. (SI Figure 1).

### Creating center-specific, de novo model validation test sets (DNMV1 and DNMV2) to enable comparison of MLCS2 and SART models

We adapted each of the 6 center-specific MLCS2 test sets to a center-specific, de novo model validation test set (DNMV1) that allows validation of MLCS2 and SART models for each center. This adaptation was performed by limiting the test set to first IVF cycles started in 2013-2022; age under 40 (required by MLCS model); IVF cycles having BMI value and yes/no for male factor, ovulatory disorder, PCOS, uterine factor and unexplained infertility diagnoses (required by SART model); yes/no for full term birth(s). The 6 DNMV1 test sets together comprised 4,645 first IVF cycles.

For the purpose of MLCS2 and SART model comparison, for each center, de novo MLCS2 model validation was performed by obtaining MLCS2 model responses for each center’s DNMV1 test set. Similarly, each center’s own SART model responses (aka predictions) were obtained by using pre-treatment model formulae, with and without AMH predictor, reportedly used to support the online SART calculator, as provided in the supplement of McLernon et al., 2022 (26). Other than to confirm the accurate implementation of those formulae using a few test cases specifically for this study, we did not interact with or use the online SART calculator website for this research study.

In the context of IVF live birth prediction models, model training required labeling each IVF cycle has having the binary outcomes “live birth” versus “no live birth”. One intentional design difference between MLCS and SART models is that in MLCS2, the models are trained with live births and clinical ongoing pregnancies labeled as “live birth”, whereas the SART model labeled live births as “live birth” and clinical ongoing pregnancies as “no live birth” (26).

Since the impact of this intentional design difference was not known, we reasoned that we should create test sets void of IVF cycles with clinical ongoing pregnancies, so that we could be sure any model comparison results are not simply due to those cycles having differential outcome labeling between MLCS and SART models. Therefore, we created a second set of 6 independent test sets (DNMV2) by removing the ∼4.8% of IVF cycles with clinical ongoing pregnancies from the DNMV1 test sets.

### Statistical analyses involving model metrics used to compare MLCS2 and SART models

The MLCS2 and SART models were compared using six independent, center-specific DNMV1 test sets for Precision, Recall, Precision Recall AUC (PR AUC), and F1 scores to allow for reporting of median and interquartile range of the model metrics, whereas one aggregate DNMV1 test set was used to compare MLCS and SART models for continuous net reclassification improvement (NRI). The entire process of model comparisons -- six independent center-specific DNMV test sets and one aggregate DNMV test set -- were repeated using DNMV2 test set (24, 33, 53–55, 58). Model metrics were reported using median and interquartile range (IQR) across 6 centers. Wilcoxon signed-rank test, allowing for non-parametric paired-testing, was used to compare MLCS2 and SART model metrics paired by center (58).

Reclassification measured the percentage of cases having different live birth predictions from the two models (59, 60). To provide clinical context, we created a 4x4 reclassification table for the DNMV1 test set to show the concordance and discordance of LBPs made by MLCS2 and SART models. Patients are placed into one of 16 groups based on their LBPs as computed by MLCS2 and SART (MLCS2-LBPs and SART-LBPs, respectively), using 4 arbitrarily defined, yet clinically intuitive prognostic categories based on LBPs: Low LBP (LBP <25%), Medium LBP (LBP 25-49.9%), High LBP (LBP 50-74.9%) and Very High LBP (LBP ≥75%), (Table 4). Patient groups are concordant if SART and MLCS2 models placed the patients into the same prognostic category (colored green in Table 4); patient groups are discordant if SART and MLCS2 models placed patients into different prognostic categories (colored blue or peach in Table 4). The same procedure was applied to the DNMV2 test set.

Further, an alternative method, continuous net reclassification improvement (NRI) was used to measure the likelihood of re-assigning a higher or lower IVF live birth probability with MLCS2 compared to SART models (60). The age-based live birth rates for each clinic stated in the finalized 2020 SART National Summary were used as age control models because practically, that is the number that providers and patients would use if they were not using any prediction models (61).

The EQUATOR Reporting Guidelines including “TRIPOD + AI statement: updated guidance for reporting clinical prediction models that use regression or machine learning methods” were followed (62, 63).

## Data Availability

The anonymized data sets containing clinical data are not available for sharing with other researchers because they are the property of each collaborating center.

## Code Availability

The code for computing model metrics and comparisons of model metrics can be shared under a confidentiality agreement for research purposes only. Code from the data pre-processing, processing and model pipelines are proprietary and can be shared via licensing. However, we would be open to supporting or collaborating with other researchers to accelerate your research.

## Acknowledgements

The authors thank the following individuals for their assistance, editing, advisory, insightful comments and contributions to the present research: Faith Ripley, BS, CPC (PREG); Patrick McCarthy, MBA (Poma Fertility); Amanda McCarthy, MBA (Poma Fertility); Brijinder S. Minhas, PhD, HCLD, MBA (NewLIFE); Wing H. Wong, PhD (Advisor); Vincent Kim, B.Sc. (Univfy Inc.); Marco Menabrito, MD (Univfy Inc.); Anjali Wignarajah, M.Sc. (Univfy Inc.); Candice Ortego (Univfy Inc.); Athena T. R. Wu (editing).

## Authorship Contribution

ETN and MWMY contributed to the original study conceptualization, design, methodology, preparation of data subsets for analyses, statistical analyses, data interpretation and visualization; writing of the original manuscript drafts and revisions. ETN, TS, XC contributed to code development, data processing, modeling, and methodologies related to those processes. ETN, MWMY, TS, XC contributed to curation of processed data. MGR, LAG, JEN, JFP, BAR, MO, JG, RB, LN, GN, JA contributed to conceptualization of study, data collection, curation of data, interpretation of data processing and modeling results, review and revision of manuscript. All authors approved submission of the manuscript for publication.

## Ethics Declaration

M Yao is employed as CEO by Univfy Inc. and is board director, shareholder and stock optionee of Univfy; she is inventor or co-inventor on Univfy’s issued and pending patents and receives payment from patent licensor (Stanford University). ET Nguyen, T Swanson, X Chen are employed by and received stock options from Univfy Inc. M Retzloff performs paid consulting work as Nexplanon trainer for Organon and is Treasurer for the Society for Reproductive Technology (SART).

## Supplementary Information

Supplementary Figure 1.

## Abbreviations

MLCS2: machine learning, center-specific model version 2
SART: Society of Assisted Reproductive Technologies
AMH: serum anti-mullerian hormone level
BMI: body mass index
Day 3 FSH: serum day 3 follicular stimulating hormone level
PCOS: polycystic ovarian syndrome
PCO: polycystic ovaries
IVF: in vitro fertilization

---

**SI Figure 1.**
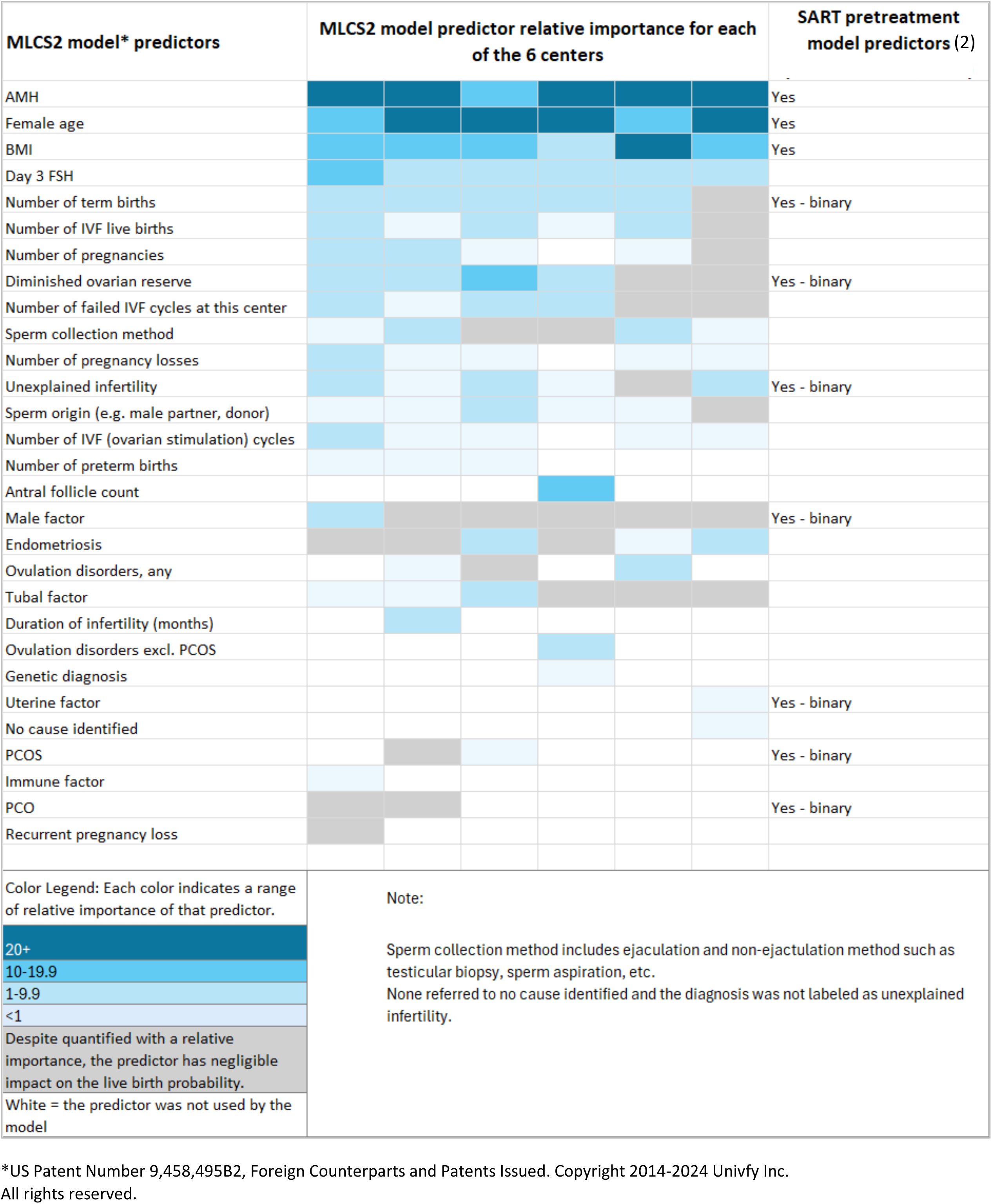
MLCS2 model predictors and their relative importance, cross-referenced with SART pretreatment model predictors*. (**1–3**) The model predictors used by each center’s MLCS2 model and their relative importance vary across centers, despite drawing from a similar set of clinical variables. The SART pretreatment model predictors were either also used by MLCS2 models or were determined to have no non-redundant predictive impact based on other predictors used by the MLCS2 models. The MLCS2 models used additional predictors not used by the SART model even though they were recorded in SART-CORS. See Yao et al, 2024 for more in-depth discussion of model predictors and relative importance (1).

