## Supplemental Figure 1 for "Predicting IVF live birth probabilities using machine learning, center-specific models: validation results and potential benefits over national registry-based models"

**SI Figure 1. MLCS2 model predictors and their relative importance, cross-referenced with SART pretreatment model predictors*. (1-3)** The model predictors used by each center's MLCS2 model and their relative importance vary across centers, despite drawing from a similar set of clinical variables. The SART pretreatment model predictors were either also used by MLCS2 models or were determined to have no non-redundant predictive impact based on other predictors used by the MLCS2 models. The MLCS2 models used additional predictors not used by the SART model even though they were recorded in SART-CORS. See Yao et al, 2024 for more in-depth discussion of model predictors and relative importance (1).

Abbreviations.
MLCS2 = machine learning, center-specific model version 2
SART = Society of Assisted Reproductive Technologies
AMH = serum anti-mullerian hormone level
BMI = body mass index
Day 3 FSH = serum day 3 follicular stimulating hormone level
PCOS = polycystic ovarian syndrome
PCO = polycystic ovaries
IVF = in vitro fertilization

| **MLCS2 model* predictors** | **MLCS2 model predictor relative importance for each of the 6 centers** | | | | | | **SART pretreatment model predictors** (2) |
|---|---|---|---|---|---|---|---|
| AMH | | | | | | | Yes |
| Female age | | | | | | | Yes |
| BMI | | | | | | | Yes |
| Day 3 FSH | | | | | | | |
| Number of term births | | | | | | | Yes - binary |
| Number of IVF live births | | | | | | | |
| Number of pregnancies | | | | | | | |
| Diminished ovarian reserve | | | | | | | Yes - binary |
| Number of failed IVF cycles at this center | | | | | | | |
| Sperm collection method | | | | | | | |
| Number of pregnancy losses | | | | | | | |
| Unexplained infertility | | | | | | | Yes - binary |
| Sperm origin (e.g. male partner, donor) | | | | | | | |
| Number of IVF (ovarian stimulation) cycles | | | | | | | |
| Number of preterm births | | | | | | | |
| Antral follicle count | | | | | | | |
| Male factor | | | | | | | Yes - binary |
| Endometriosis | | | | | | | |
| Ovulation disorders, any | | | | | | | |
| Tubal factor | | | | | | | |
| Duration of infertility (months) | | | | | | | |
| Ovulation disorders excl. PCOS | | | | | | | |
| Genetic diagnosis | | | | | | | |
| Uterine factor | | | | | | | Yes - binary |
| No cause identified | | | | | | | |
| PCOS | | | | | | | Yes - binary |
| Immune factor | | | | | | | |
| PCO | | | | | | | Yes - binary |
| Recurrent pregnancy loss | | | | | | | |

Color Legend: Each color indicates a range of relative importance of that predictor.
- 20+
- 10-19.9
- 1-9.9
- <1
- Despite quantified with a relative importance, the predictor has negligible impact on the live birth probability.
- White = the predictor was not used by the model

Note:

Sperm collection method includes ejaculation and non-ejactulation method such as testicular biopsy, sperm aspiration, etc.
None referred to no cause identified and the diagnosis was not labeled as unexplained infertility.

References.


1. Yao MWM, J Jenkins, Nguyen ET, Swanson T, Menabrito M. Patient-centric IVF prognostics counseling using machine learning for the pragmatist. Semin Repro Med 2024, *accepted*.

2. McLernon DJ, Raja EA, Toner JP, Baker VL, Doody KJ, Seifer DB, Sparks AE, Wantman E, Lin PC, Bhattacharya S, Van Voorhis BJ. Predicting personalized cumulative live birth following in vitro fertilization. Fertil Steril. 2022 Feb;117(2):326-338. doi: 10.1016/j.fertnstert.2021.09.015. Epub 2021 Oct 19. PMID: 34674824.

3. Curchoe CL, Tarafdar O, Aquilina MC, Seifer DB. SART CORS IVF registry: looking to the past to shape future perspectives. J Assist Reprod Genet. 2022 Nov;39(11):2607-2616. doi: 10.1007/s10815-022-02634-6. Epub 2022 Oct 21. PMID: 36269502; PMCID: PMC9722991.



*US Patent Number 9,458,495B2, Foreign Counterparts and Patents Issued. Copyright 2014-2024 Univfy Inc. All rights reserved.
